# Network Profile: Improving Response to Malaria in the Amazon through Identification of Inter-Community Networks and Human Mobility in Border Regions of Ecuador, Peru, and Brazil

**DOI:** 10.1101/2023.11.29.23299202

**Authors:** Mark M. Janko, Andrea L. Araujo, Edson J. Ascencio, Gilvan R. Guedes, Luis E. Vasco, Reinaldo A. Santos, Camila P. Damasceno, Perla G. Medrano, Pamela R. Chacón-Uscamaita, Annika K. Gunderson, Sara O’Malley, Prakrut H. Kansara, Manuel B. Narvaez, Carolina S. Coombes, Francesco Pizzitutti, Gabriela Salmon-Mulanovich, Benjamin F. Zaitchik, Carlos F. Mena, Andres G. Lescano, Alisson F. Barbieri, William K. Pan

## Abstract

**Objectives:** Understanding human mobility’s role on malaria transmission is critical to successful control and elimination. However, common approaches to measuring mobility are ill-equipped for remote regions such as the Amazon. This study develops a network survey to quantify the effect of community connectivity and mobility on malaria transmission.

**Design:** A community-level network survey

**Setting:** We collect data on community connectivity along three river systems in the Amazon basin: the Pastaza river corridor spanning the Ecuador-Peru border; and the Amazon and Javari river corridors spanning the Brazil-Peru border.

**Participants:** We interviewed key informants in Brazil, Ecuador, and Peru, including from indigenous communities: Shuar, Achuar, Shiwiar, Kichwa, Ticuna, and Yagua. Key informants are at least 18 years of age and are considered community leaders.

**Primary outcome:** Weekly, community-level malaria incidence during the study period.

**Methods:** We measure community connectivity across the study area using a respondent driven sampling design. Forty-five communities were initially selected: 10 in Brazil, 10 in Ecuador, and 25 in Peru. Participants were recruited in each initial node and administered a survey to obtain data on each community’s mobility patterns. Survey responses were ranked and the 2-3 most connected communities were then selected and surveyed. This process was repeated for a third round of data collection. Community network matrices will be linked with eadch country’s malaria surveillance system to test the effects of mobility on disease risk.

**Findings:** To date, 586 key informants were surveyed from 126 communities along the Pastaza river corridor. Data collection along the Amazon and Javari river corridors is ongoing. Initial results indicate that network sampling is a superior method to delineate migration flows between communities.

**Conclusions:** Our study provides measures of mobility and connectivity in rural settings where traditional approaches are insufficient, and will allow us to understand mobility’s effect on malaria transmission.

**Strengths and Limitations:**

1. Strength: Community networks are unmeasured in rural areas of the Amazon, but have been shown to capture human mobility in other regions of the world.
2. Strength: Our design captures social, economic, and human wellbeing connectivity and migration in key indigenous communities along the Peru-Ecuador border as well as in the most important confluence for the Amazon River located in the Brazil-Peru-Colombia tri-country intersection.
3. Strength: Our design quantifies cross-border human mobility between communities, as well as the magnitude, timing, duration, and reason for mobility, which provides actionable information for malaria control and elimination programs in the region
4. Limitation: Migration decisions occur at individual and household levels that are coupled with environmental change and seasonality, meaning that our measures of community mobility may not be stable over time and we may be subject to ecological fallacy by inferring individual risk from community networks.
5. Limitation: Our study relies on passive surveillance to test the community network/human mobility link with malaria. However, there exist cases that are asymptomatic, unreported (i.e., treated with traditional medicines), or that occur in our community network but are reported elsewhere. The extent of these cases can significantly increase uncertainty.

**Funding:** This work was supported by the US National Institutes of Health (R01 AI51056; William K. Pan, PI) and by a grant from the Duke Climate and Health Initiative (William Pan, PI). PRC-U was supported by CONCYTEC through the PROCIENCIA program under the call entitled “Science, Technology and Innovation Thesis and Internships” according to the contract PE501081617-2022. AGL, CSC, EJA and PRC-U were sponsored by Emerge, the Emerging Diseases Epidemiology Research Training grant D43 TW007393 awarded by the Fogarty International Center of the US National Institutes of Health.

**Competing Interests:** We declare no conflicts

## 1. Introduction

Despite considerable progress in reducing the burden of malaria since 2000, progress has stalled in recent years, and malaria still kills over 600,000 people per year (1). Nevertheless, different initiatives around the world are underway with the hope of eventually eliminating the disease, including in the Americas, which is characterized by relatively low, unstable transmission (2). An important challenge to elimination in the Americas and elsewhere is our insufficient understanding of human mobility and its effects on transmission. Human mobility can undermine malaria control strategies in three important ways. First, human mobility among malaria-infected individuals causes the continuous flow of parasites into and out of communities that are working to reduce and potentially eliminate transmission (3,4). Second, mobility contributes to the diffusion of novel malaria strains that can be drug-or diagnostic-resistant, which can render front-line control measures ineffective (3,5). Third, in the Amazon, human mobility drives environmental change, which can fundamentally alter vector ecology and the epidemiology of transmission (6,7). Understanding human mobility and developing migration-relevant interventions is critical to successful malaria elimination, particularly in the Amazon, where malaria risk varies from hyper-endemic to seasonally unstable, making transmission highly sensitive to changes in malaria control investments (8).

Although several studies highlight the importance of migration and mobility on malaria transmission, few (if any) control programs integrate knowledge of local, regional or international human migration into control strategies (9–12). This is particularly problematic in the Amazon, where human mobility patterns are heterogeneous—involving short- and long-term movements that include circular migration, commuting and permanent out-migration over different temporal scales—all of which contribute to seasonal and sub-seasonal malaria patterns (13–15). To be informative for malaria control, information on human mobility must provide information on three key features: (1) the timing and spatial extent of movement; (2) origin and destination; and (3) push/pull factors of movement. These features are needed for control programs to determine when and where to deploy interventions, as well as to develop interventions relevant to the migrant. Unfortunately, existing methods measure human migration and link mobility to malaria are limited in their ability to address these needs. For example, mobile phone data has been linked to malaria surveillance data to quantify the effect of mobility on transmission, but it does not address why people move (16–21). In the Amazon, residents travel for a variety of reasons, including to work in extractive or other environmentally-linked trades implicated in malaria transmission, such as logging, mining, fishing, or hunting (8,12,15,22–24). Others may travel to buy and sell goods, receive healthcare, participate in social or political activities, or send children to school. Additionally, in rural regions of the Amazon where malaria transmission is highest, cell phone coverage is sparse or non-existent, meaning even basic measures of mobility are unavailable across the vast majority of the region (25).

Other work focusing on human movement, including participatory mapping and GPS tracking studies, have demonstrated explicit links between mobility, seasonal and environmental variability, and malaria transmission at small (e.g. a few kilometers) spatial and weekly temporal scales (12,22,26,27). However, these studies generally provide limited inferences for malaria control as they focus on individual-level movements within a community, which may be different from general patterns of mobility between communities, thereby limiting the utility for community-based malaria control.

Conversely, studies focusing on between-community mobility have relied on a variety of modeling approaches, including gravity models, radiation models, and metapopulation models, to name a few (28–31). Gravity models assume that movement between two locations increases as a function of their population sizes, and decreases with distance between them. These measures have been linked to malaria surveillance data to quantify importation rates between regions, but this approach was found to be inferior to metapopulation models that leveraged human mobility from cell phone networks (28). Conversely, other modeling efforts designed to understand malaria importation between locations using radiation models, which assume that travelers are absorbed into surrounding communities as a function of the distance from their origin community and the population size within a radius of the distance travelled. One comparison made with respect to understanding malaria diffusion noted that gravity models are better suited for travel to large population centers, whereas the radiation model provided better fits to more localized travel (29). Nevertheless, model performance varied according to different groups of travelers (e.g. young workers, mothers with small children), highlighting the need to collect data on different travel groups to understand their mobility patterns and how those might contribute to malaria diffusion in different ways (29). This need is reinforced by other work, which has shown that various classes of models to understand the impact of mobility on malaria are highly sensitive to the choice of human movement model (31).

Finally, studies on mobility and malaria have been enhanced by molecular and genomic surveillance, which allow for the quantification of how parasite populations are connected over space and time (32–37). However, these studies also possess important limitations. First, human mobility is not directly measured, but is inferred based on genetic similarities between parasite populations in different locations. Second, similar to cell-phone based or modeling studies, these studies cannot identify the human migration processes that drive malaria diffusion. Third, genomic studies have often been conducted at large spatial and temporal scales, tracking parasite movement between countries or provinces over several years (20,38– 40), masking seasonal variability in mobility patterns at more localized spatial and temporal scales. This is particularly important in low-transmission settings, where spatially focal and seasonal surveillance efforts are needed for elimination (9). Finally, the genomic sequencing technologies needed to understand localized patterns of parasite diffusion are often cost-prohibitive.

Therefore, better measures of human mobility that characterize the drivers, timing, spatial extent, and magnitude are needed to guide control. To address this knowledge gap, we designed a study to evaluate human mobility patterns and drivers across almost 300 communities near the Ecuador-Peru and Peru-Brazil border regions, and how these patterns contribute to seasonal and sub-seasonal patterns of malaria. We develop a novel community-based social network analysis that evaluates migration networks between communities to test the primary hypothesis that community relational ties (social, economic, transport) are correlated with spatio-temporal malaria patterns. These data will also be incorporated into Agent-Based Models (ABMs) evaluating different intervention scenarios to guide malaria control in border communities. Finally, we compile a comprehensive dataset spanning infrastructure, demographic, land use/land cover, and other environmental data to test how observed patterns of human mobility may relate to region-wide malaria patterns across all of the Ecuadorian, Peruvian, and Brazilian Amazon.

This paper is organized as follows. Section 2 introduces the study settings. Section 3 describes the community network sampling design and pilot tests in Brazil, Ecuador, and Peru. Section 4 details the analysis plan.

## 2. Study Setting

This study takes place along three river corridors in the northwestern Amazon: the Pastaza river, the Amazon (or Amazonas/Solimões) River, and the Yavari (or Javari) River (Figure 1). Regions defined by these three river systems vary considerably in terms of malaria transmission intensity, ecology, demography, and economy.

### 2.1 Pastaza River Corridor

The region comprising the Pastaza River corridor makes up part of the southeastern Ecuadorian and northwestern Peruvian Amazon (Figure 1). The river runs for 710 kilometers, beginning with headwaters from the Ecuadorian Andes and ending when its mouth empties into the Marañon River in Peru. Our study area comprises the Pastaza River corridor from Puyo, Ecuador to the Marañon River in Peru. Approximately half of the study area is in Ecuador and half in Peru. The area has a relatively flat terrain, with rainfall throughout the year, the heaviest occurring from November to May. Maximum temperature varies from 24 to 35 °C and the minimum temperature varies from 20 to 25 °C, with the October through January being the warmer months. Land use and land cover are mostly comprised of forest, with a minor proportion of agricultural land, shrub and grass vegetation, and permanent and seasonal wetlands. The study area in Ecuador comprises 13 parishes with a sparse population of 2,850 (2010 Census). The study area in Peru comprises 5 districts (Andoas, Trompeteros, Pastaza, Lagunas and Barranca) with a sparse population of 63,929 in 2022. The study area passes through the Achuar indigenous territory, which is home to an estimated 18,500 Achuar people who live within an area of approximately 12,000 square kilometers. In both Ecuador and Peru, the most populated and urbanized communities are the main commercial points to buy and sell products, go to school, and obtain healthcare. In Peru, economic activity in native and rural communities mostly consists of agriculture and fishing, as well as selling their products in the main cities. In the Ecuadorian side, the main economic activities are tourism, agriculture, wood extraction, and handicrafts.

This river corridor is a hotspot for malaria transmission in both Ecuador and Peru. In Ecuador, malaria incidence was 39 cases per 1,000 inhabitants in 2021 (the most recent year for which data are available), and varied across parishes from 0 to 290 cases per 1,000 residents. Across the border in Peru, malaria incidence in the study area was 230 cases per 1,000 inhabitants in 2022, and varied across communities from 140 to 275 cases per 1,000 inhabitants.

### 2.2. Amazon (Solimões) and Javari River Corridors

The Amazon and Javari River corridors correspond to the northwestern-most part of the Brazilian Amazon, and the easternmost part of the northern Peruvian Amazon (Figure 1). The area has a relatively flat terrain and a regime of heavy rainfall. The study area comprises six municipalities in Brazil (Amaturá, São Paulo de Olivença, Santo Antônio do Içá, Tabatinga, Atalaia do Norte, and Benjamin Constant), and three districts in Peru (Javari, San Pablo, and Ramon Castilla). The region in both Brazil and Peru is sparsely populated, like most of the Amazon region. In Brazil, the region is home to an estimated 207,903 inhabitants as of 2021. In Peru, the region was home to 74,118 inhabitants as of 2020. The cities and communities are largely connected via transit along rivers, with the Javari and Solimões (Amazon) Rivers as the main waterways.

In 2021, malaria incidence in Peru in this region was 1.4 cases per 1,000 residents, which varied across districts from 0.5 to 30 cases per 1,000 residents. Across the border in Brazil, malaria incidence was 8.6 cases per 1,000 residents, which varied across municipalities from 0.1 to 100 cases per 1,000 residents.

## 3. Study Design

### 3.1 Respondent Driven Sampling protocol

As noted in the introduction, the motivations for mobility are diverse, including buying and selling goods, accessing health services, studying, obtaining services, participating in social, religious, or political activities, looking for work, or visiting family, to name a few. The intensity of mobility may vary across motivations for travel, and seasonally. Further, there are important differences in exposure beyond seasonality, such as length of stay at a given destination and its ecological suitability for malaria transmission. To capture these dimensions of human mobility, we leverage a respondent driven sampling (RDS) design with up to three waves of data collection, in which the initial wave of data collection produces candidate communities for the second wave of data collection, which, in turn, produces candidate communities for the third and final round of data collection. The *community* is therefore the primary unit of analysis.

To implement this study design, the first wave of data collection required the selection of initial node communities. We selected these initial node communities based on malaria incidence rates (low, medium, high), demographics (e.g. indigenous communities or not), and geography (e.g. distance from international border). A total of 21 initial nodes were selected along the Pastaza river corridor (10 in Ecuador, 11 in Peru), while 20 initial nodes were selected along the Amazon and Javari corridors (10 in Brazil, 10 in Peru) (Figure 1; Table 1). An additional three potential initial nodes were added for Brazil since some initial nodes may only be connected to indigenous communities in protected areas, which cannot be sampled without first seeking approval from the Fundação Nacional dos Povos Indigenas (FUNAI). Additionally, the Javari river is sparsely populated, and some initial nodes may exhibit little to no connectivity, and so additional nodes are included in the event that a given node is not connected to any other eligible communities. Therefore, the 10 final initial nodes in Brazil are selected as fieldwork evolves.

**Table 1.** Characteristics of initial nodes across the Javari, Amazon, and Pastaza river systems.

| Country | Watershed | N | Malaria Risk* |  |  | Health Center | Indigenous |
| --- | --- | --- | --- | --- | --- | --- | --- |
|  |  |  | High | Medium | Low |  |  |
| Brazil** | Javari | 9 | 2 | 4 | 3*** | 0 | 0 |
|  | Amazon | 4 | 0 | 0 | 4 | 1 | 0 |
| Peru | Amazon | 8 | 3 | 0 | 5 | 4 | 4 |
|  | Yavari | 2 | 2 | 0 | 0 | 2 | 0 |
|  | Pastaza | 11 | 4 | 0 | 7* | 4 | 11 |
| Ecuador | Pastaza | 10 | 8 | 2 | 0 | 5 | 10 |

\* Risk categories are country-specific: Malaria incidence categories in Peru and Ecuador were defined as high or low based on whether or not they fell above or below the median cumulative incidence rate. In Peru, the median was 343 cases/1,000 at risk, based on data from 2009 – 2020. In Ecuador, the median was 427 cases/1,000 at risk, based on data from 2008-2020. In Brazil, High was categorized as >50 cases/1,000 at risk; Medium was 10-49 cases/1,000 at risk, and Low was <10 cases/1,000 at risk.
\*\* Three nodes are included as potential initial nodes to allow for nodes to be replaced if they are only connected to indigenous communities in protected areas, which are currently not eligible for enrollment in the study.
\*\*\* There was one community without malaria reported (i.e., missing) assigned as Low Risk. The dotted line separates the Brazil-Peru site from the Ecuador-Peru site

Due to the RDS design, the selection of waves two and three of data collection depends on responses by study participants within each community. To elicit these responses, we designed a survey about general mobility patterns and recruit key informants within each community. Key informants are male and female community leaders (religious, political, business, etc.) who are at least 18 years of age. Key informants are asked to name up to six communities where members go for different reasons (outflow) and up to six communities where residents of other communities come from for different reasons (inflow). They are further asked to identify the number of people who travel to or from each named community in a given year, the timing of peak travel (rainy season, dry season, year-round), the length of travel, the average number of nights stayed in a community for a typical visit, and whether or not the named community is perceived to be a source of malaria importation. Additional data about the presence of different infrastructure within the community are collected (e.g. schools, health care facilities, a central market, church, electricity, cell phone coverage, potable water, etc.). Finally, in communities where there is a community health worker or other health infrastructure (e.g. a health post), a local health professional is also interviewed about residents’ travel related to health care services and the availability of services within the community. Data about the following services were collected: family planning, maternal health care, pediatric health care, malaria diagnostics, malaria treatment, services to treat minor injuries/illnesses, surgical services, or dental services. Finally, health professionals are asked about the presence of different personnel in the community, including physicians, nurses, midwives, or microscopists. All surveys were translated into Spanish, Portuguese, and Achuar, as necessary, and are available in the accompanying supplement.

Upon interviewing participants in a given initial node community, the survey team then ranks connectivity between the initial node and the communities identified in the surveys. Connectivity is ranked in two different ways. First, connectivity is ranked overall, as follows:

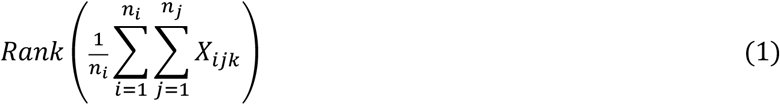

Where *X*_*ijk*_ is the total number of people key informant *i* reports traveling for reason *j* between the current community and community *k*. Summing over all reasons *n*_*j*_ and across all key informants *n*_*i*_ yields the total number of people traveling for all reasons of travel between the initial node and identified community. We then normalize by the number of key informants (*n*_*i*_) interviewed in community *k* to account for any variability in the number of key informants recruited across communities. Finally, we rank the set of *k* communities generated and select the community with the greatest overall connectivity.

Second, connectivity is ranked by reason of travel by summing the total number of people traveling for a given reason and dividing by the number of key informants recruited. Two to three Wave 2 communities are then enrolled per initial node. The first of these communities is selected based on the greatest overall connectivity to the initial node, while the remaining community or communities are selected based on the top two reasons of travel (excluding the first selected community in the event that it also reports the top reason of travel). This is done to ensure that communities are selected across the diverse reasons of travel in the region. The same procedure is followed for Wave 3 communities.

### 3.2 Pilot Test

Given the need to rapidly assess community connectivity patterns to inform subsequent waves of data collection across three countries in difficult field conditions with travel across long distances, we conducted pilot tests to inform the feasibility of this design and make any needed modifications. We conducted pilot tests in Peru (June 2022), Ecuador (July 2022), and Brazil (July 2022) in eight of the initial nodes selected (two in Peru; four in Ecuador; two in Brazil). The objectives of the pilot tests were to: 1) compare responses generated via key informants who are organized into focus groups versus responses generated by key informants interviewed individually; 2) assess the number of key informants or size of focus group necessary to reach a consensus view of connectivity; 3) compare the time needed to conduct each survey using REDCap on both tablets and mobile phones; and 4) introduce the study locally and identify potential local surveyors whose knowledge of the study areas would maximize study success. In each community, we conducted between two and six key informant interviews. In Peru and Ecuador, we conducted focus groups comprised of both males and females, as well as male- and female-only focus groups.

Findings indicated that paper-based surveys with at least 4 key informant interviews were superior to focus groups and REDCap data entry. Paper-based surveys with key informants were easier to administer, requiring less time to coordinate multiple participant schedules in focus groups, and less time to record responses (20 minutes versus >1 hour for REDCap), particularly among women, who often had childcare responsibilities. We observed little variability in responses between data provided by key informants versus focus groups, and little variability across key informants, indicating that survey questions about mobility were well understood and we did not require a large number of informants per community.

Another important observation was the report of travel to urbanized areas that do not involve overnight stays, making them destinations with little to no malaria risk (since malaria is transmitted from dusk to dawn). Thus, these types of communities (i.e., Benjamin Constant in Brazil or Caballococha in Peru) are excluded from subsequent waves. In addition, we recognized major obstacles to visiting and enrolling communities due to their remote locations, particularly as we extend the network beyond the 2^nd^ wave. Thus, to ensure data collection is conducted in a safe, timely and economical manner, survey teams are empowered to substitute tertiary communities with those that are more accessible, if needed.

### 3.3 Data collection to date

#### Pastaza River corridor

Fieldwork occurs separately in the Pastaza River corridor spanning the Ecuador-Peru border and the Amazon/Javari River corridors spanning the Brazil-Peru border. Fieldwork along the Pastaza River began in October 2022 with the survey administered in the 10 Ecuadorian and 11 Peruvian initial nodes. This fieldwork was completed in November 2022. From these 21 initial nodes, a total of 57 unique communities were identified with connectivity across eight reasons for movement: buying or selling goods, education, seeking healthcare, participating in political activities, participating in religious activities, participating in social activities, seeking work, and other (which included fishing, using government services, and accessing the internet) (Figure 2). The magnitude of travel varied from as few as a single person moving between communities to a maximum of 2,400 moving for work, which we observed between the initial node of Kumai and the provincial capital of Puyo, which sits at the northern extent of the study area (Figure 1).

Because of the Christmas and New Years holidays, data collection for Waves 2 and 3 could not begin until 2023. Therefore, in the time between the first and second wave of data collection, we selected Wave 2 communities based on the protocol identified above, such that the selection would not need to be done in the field. This allowed cross-border referrals to be ranked and selected more easily, since the ability of the Ecuadorian and Peruvian field teams to communicate in the region is limited. However, Wave 3 community selection was done in the field. Wave 2 data collection began in the Ecuador segment of the Pastaza corridor in March 2023 and all three waves of data collection were completed in May 2023. No referrals for Wave 3 communities from Ecuador included Peruvian communities that had not already been surveyed in Wave 1 in Peru. Across all three waves of data collection, a total of 126 communities were surveyed (76 in Ecuador; 50 in Peru), with data collected from 586 key informants (319 in Ecuador; 267 in Peru), which yielded an average of 4.7 key informants per community. Key informants had a median age of 36 years in Ecuador (minimum age 18, maximum age 78) and 40 years in Peru (19 – 80). A total of 208 key informants were female, and 378 were male. Data collection in Peru and Brazil is ongoing. As data is collected, it is entered into a REDCap database hosted by the Universidad Peruana Cayetano Heredia (Ecuador and Peru) and Universidade Federal Minas Gerais (Brazil) for subsequent analyses (41,42).

#### Amazon River corridor

Data collection began in the Brazil side of the Amazon corridor in May 2023, while the Peru side is scheduled to begin in September 2023. The staggered start dates allow for the final selection of the Brazilian initial nodes, and for advanced referrals to the Peru team for inclusion in the Peru fieldwork along the Amazon river. To date, data collection has been completed for two initial nodes and the Wave 2 and 3 communities referred to from them, with a total of 14 communities being enrolled. Interviews across each initial node and the referral communities took a total of 4 days, including travel. The fieldwork along the Amazon river corridor is scheduled to finish in late 2023.

#### Javari River corridor

Data collection along the Javari River corridor is scheduled to begin in July 2023, starting in Brazil followed by Peru. The staggered start dates will allow for Brazilian connections in Peru to be incorporated in the Peru fieldwork. Data collection in Brazil will similarly span the length of the Peru data collection, such that any referrals to Brazilian communities from Peru can be surveyed.

## 4. Analysis Plan

### 4.1 Linking network data to surveillance, environmental, and infrastructure data

When survey work is completed and data are entered into a database, we will construct community connectivity matrices, one for each identified reason of travel. Each community network matrix *W*_*j*_ is an *n* × *m*_*j*_ matrix where each row corresponds to a community visited and each column corresponds to a community identified through the network survey described above. Each element of the matrix has the total connectivity between community *i* and *j*. We allow for a greater number of columns *m* than rows *n* since it is likely that more communities will be referred to than visited. The subscript *j* denotes the columns formed from communities identified for a given reason of travel.

For each community surveyed and its identified connections, we then link this to the malaria surveillance data available from each country’s surveillance system. Our ability to link these data is due to all three countries (as well as the broader Americas region) having similar disease surveillance systems and protocols, which were developed with support from the Pan American Health Organization (43). Specifically, microscopy-confirmed case counts (by species: *P. falciparum* and *P. vivax*) are reported at the community level for each epidemiological week across all three countries. Given that the survey period comprises roughly the period spanning October 2022 to August 2023, we will conduct our primary analysis by linking malaria surveillance data from this period to the observed network. Further, because we collect data on the seasonality of travel, we allow for the entries in each *W*_*j*_ to vary across the rainy and dry seasons, as needed.

Despite having similar surveillance systems, linking the malaria surveillance data also presents challenges, including the fact that malaria cases are reported in multiple different ways. For example, malaria cases are reported either in the community where the patient resides, in the community where the diagnosis is made (typically a community with either a health post or a community health worker), or where the patient reports recent travel (i.e. the case is suspected to be imported). We address this challenge as follows. Where possible, suspected imported cases will be linked to the probable community of infection. Second, for those not reporting recent travel, we will assign cases to the community in which residents live. Finally, remaining cases will be assigned to the community where they were diagnosed. We will conduct sensitivity analyses to determine if our case assignment improves the model fit (section 4.2) over simply assigning cases to where they are diagnosed.

In addition to the network data, our team has developed a Land Data Assimilation System (LDAS) that provides hydrometeorological data (e.g. temperature, precipitation, surface runoff, soil moisture content, surface temperature) at the same spatial and temporal scales as the malaria surveillance system across the study region (24,44,45). Further, we conducted a web scraping exercise to obtain publicly available demographic (e.g. census), infrastructure (e.g. roads, schools, health centers, etc.), and land cover data. LDAS, land cover, and community-specific web-scraping data (e.g. whether or not a given community has a school, has internet access, as well as demographic data) is linked to the malaria surveillance and network data using either Geographic Information System (GIS) tools, or by merging based on either the administrative codes for levels two and three, or the community-level code for Brazil, Ecuador, and Peru. We include a further discussion of the web-scraping activity, as well as a comprehensive list of the hydrometeorological, land cover, demographic, and infrastructure data in a supplementary appendix.

### 4.2 Statistical analysis

To test our hypothesis that community relational ties are directly proportional to spatio-temporal malaria patterns, regardless of physical distance, we work within a Bayesian framework and fit network autocorrelation models using the following general form:

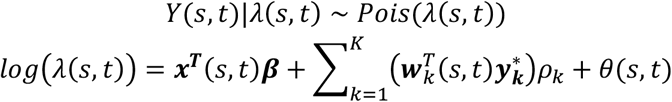

where *Y*(*s, t*) is the number of malaria cases (by species) in community *s* and epidemiological week *t*, ***x***^***T***^(*s, t*) is a vector of demographic, economic, and ecological covariates, ***β*** is a vector of regression coefficients linking these covariates to the response, ***w***^***T***^(*s, t*) is a vector of community network ties for each reason of travel *k*, while 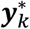 is a vector of temporally lagged malaria incidence rates in each community connected to community *s* for reason *k*. Combined, 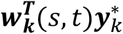 represents a measure of how malaria flows between communities and along community networks for a given reason, while the parameter *ρ*_*k*_ captures the association between network connectivity and malaria incidence. Different lags will be explored (e.g. 4 weeks vs 8 weeks). Finally, *θ*(*s, t*) represents a random effect, which can be modeled in a variety of ways, including spatially or spatio-temporally using Gaussian Process priors.

Working within a Bayesian setting makes the above model highly flexible. We will therefore consider different parameterizations for the above model. For example, *ρ*_*k*_ can be specified as a vector (***ρ***_*k*_) and allowed to vary by community, thereby allowing the effect of community network ties on malaria transmission to vary across communities (or networks, in the case where the overall total network is made up of smaller networks). The network connectivity vector ***w***^*T*^(*s, t*) can also be constructed in different ways and define network connectivity based on populations flows at different times, such as by season. Thus, this general modeling approach will allow the effects of human mobility on malaria transmission to be identified while also accounting for drivers and timing of mobility, as well as ecological, demographic, and economic conditions. All models will be fit in R using bespoke MCMC algorithms. We will stratify by malaria species, and adjust for population at risk through an offset, and consider different likelihood functions (e.g. negative binomial). We will consider stratifying by different network clusters. We will use root mean square prediction error (RMSPE) for model comparison, and base inferences on the best fitting model (for the overall network) or models (for networks).

## 5. Discussion

This network will enable the measurement of human mobility patterns, their drivers, and their relationship to key infectious disease outcomes across three river systems in the northwestern Amazon, including cross-border movements, while accounting for environmental, demographic, and other confounders. Our network focuses on remote communities of the Amazon where health care access is limited, disease incidence is both high and likely undercounted, and families are highly vulnerable to environmental, economic, and political shocks that can significantly impact well-being. In addition, our cohort was selected to specifically obtain data on indigenous communities, who typically experience the highest malaria and other health burdens. Finally, our approach has a number of advantages over other studies seeking to quantify human mobility across regions, which have relied on cell phone data, transportation networks, or inferred human mobility through genomic or molecular studies.

One potential limitation of this study is that community mobility patterns may change over time. For example, occupation-related mobility is seasonal and climate sensitive, and climate change may lead to changing mobility patterns, either creating new community linkages that do not currently exist, eliminate current linkages between communities, or otherwise changing the strength of the connections between communities. Additionally, our study relies on malaria surveillance to test the community network/migration link. There are both asymptomatic and non-reported cases that occur in the communities, as well as cases that occur within the social network but are reported outside. We attempt to capture all these external cases and assign them appropriately by residence, but some infections travel outside the network and aren’t captured. This limitation is a balance between longitudinal, prospective studies that only focus on 1-2 communities yet obtain rich infection data or retrospective case-control studies that try to quantify attributable risk of migration to malaria incidence versus the dynamic changes to malaria distribution that can be detected by this larger scale approach that captures socio-economic and demographic (migratory) structure. Finally, we did not collect data on indigenous communities located inside protected indigenous land since authorization to enter is pending from the Brazilian Indigenous Foundation (FUNAI). Once permission is granted, and if logistics and the political and conflict context changes, we may collect and add information on these communities in the near future.

## Data Availability

All data produced in the present study will be made available upon reasonable request to the authors upon completion of the study.

## Author contributions

MMJ, WKP, AFB, CFM, AGL, GRG, FP, and GSM designed the study. ALA, EJA, AFB, PGM, LEV, and MMJ developed the survey instruments. ALA, EJA, LEV, RAS, CPD, AGL, and AFB selected the initial nodes. ALA, LEV, PGM, and MMJ conducted the pilot test in Ecuador. EAJ, PGM, and MMJ conducted the pilot test in Peru. CPD and AFB conducted the pilot in Brazil. PHK, MBN, and BFZ developed the LDAS. AKG conducted web-scraping. PRCU, PGM, and SOM developed the REDCap database for the social network survey. CSC, AFB, CPD, RAS, and MMJ conducted fieldwork along the Amazon and/or Javari rivers. MMJ and WKP developed the statistical analysis plan. MMJ, ALA, EJA, GRG, AFB, and WKP wrote the manuscript. Funding was provided by NIH (R01-AI151056) to WKP. All authors reviewed and approved the final manuscript for submission.

