## Supplemental Data 1 for "Network Profile: Improving Response to Malaria in the Amazon through Identification of Inter-Community Networks and Human Mobility in Border Regions of Ecuador, Peru, and Brazil"

### Supplemental Material

### Survey Instruments

#### Supplementary Figure 1: Key Informant Inflow Survey

**INFLOW KEY INFORMANT FORM**

Interviewer Name:______________________

Interview Date: ________________________

**Reason for travel:**
1) Buy/sell goods
2) Health care
3) Social activities
4) Work
5) Visiting family
6) Political events
7) Religious events
8) Education
9) Other

**Season:**
1) Rainy
2) Dry
3) Both

**# of nights stayed:**
1) < 1 day
2) 1-2 days
3) > 2 days

**Malaria importation:**
1) Yes
2) No

| **Key Informant Characteristics** | | |
|---|---|---|
| First name | | |
| Last name | | |
| Sex | M | F |
| Age (18-100) | | |
| Ethnicity | | |
| Education | | |
| Occupation | | |
| Year they came to live in community (1900-2023) | | |

| **Data Upload to REDCap** |
|---|
| Uploaded by: |
| Date (dd/mm/yy): |
| Notes: |

| **Community Name:** ______________ | **Reason for travel** | **# of people coming in a year (0-10000)** | **Season** | **Travel time** | **# of nights stayed in a year** | **Malaria importation** | **Notes** |
|---|---|---|---|---|---|---|---|

### Supplementary Figure 2: Key Informant Outflow Survey

**OUTFLOW KEY INFORMANT FORM**

Interviewer Name:______________________

Interview Date: ______________________

**Reason for travel:**
1) Buy/sell goods
2) Health care
3) Social activities
4) Work
5) Visiting family
6) Political events
7) Religious events
8) Education
9) Other

**Season:**
1) Rainy
2) Dry
3) Both

**# of nights stayed:**
1) < 1 day
2) 1-2 days
3) > 2 days

**Malaria importation:**
1) Yes
2) No

| **Community Characteristics** | | |
|---|---|---|
| Infrastructure available: | | |
| School types: | | |
| - Primary education | Yes | No |
| - Secondary education | Yes | No |
| - Technical/vocational | Yes | No |
| Health post | Yes | No |
| Health center | Yes | No |
| Community center | Yes | No |
| Pharmacy | Yes | No |
| Central market | Yes | No |
| Church | Yes | No |
| Power grid | Yes | No |
| Cell phone coverage | Yes | No |
| Water supply | Yes | No |
| Market | Yes | No |
| **Data Upload to REDCap** | | |
| Uploaded by: | | |
| Date (dd/mm/yy): | | |
| Notes: | | |

| **Community Name:** ____________ | **Reason for travel** | **# of people coming in a year (0-10000)** | **Season** | **Travel time** | **# of nights stayed in a year** | **Malaria importation** | **Notes** |
|---|---|---|---|---|---|---|---|

### Supplementary Figure 3: Health Professionals Inflow Survey

**INFLOW HEALTH CARE PROFESSIONAL FORM**

Interviewer Name:_____________________

Interview Date: _______________________

**Reason for travel:**
1) Family planning
2) Maternal care
3) Pediatric care
4) Malaria diagnostic
5) Malaria treatment
6) Minor illness/injury
7) Surgery
8) Dental
9) Other

**Season:**
1) Rainy
2) Dry
3) Both

**# of nights stayed:**
1) < 1 day
2) 1-2 days
3) > 2 days

**Malaria importation:**
1) Yes
2) No

| **HCP Informant Characteristics** | | |
|---|---|---|
| First name | | |
| Last name | | |
| Sex | M | F |
| Age (18-100) | | |
| Ethnicity | | |
| Occupation | | |
| Year they came to live in community (1900-2023) | | |
| **Data Upload to REDCap** | | |
| Uploaded by: | | |
| Date (dd/mm/yy): | | |
| Notes: | | |

| **Community Name:** ____________ | **Reason for travel** | **# of people coming in a year (0-10000)** | **Season** | **Travel time** | **# of nights stayed in a year** | **Malaria importation** | **Notes** |
|---|---|---|---|---|---|---|---|

### Supplementary Figure 4: Health Professionals Outflow Survey

**OUTFLOW HEALTH CARE PROFESSIONAL FORM**

Interviewer Name:______________________

Interview Date: ________________________

**Reason for travel:**
1) Family planning
2) Maternal care
3) Pediatric care
4) Malaria diagnostic
5) Malaria treatment
6) Minor illness/injury
7) Surgery
8) Dental
9) Other

**Season:**
1) Rainy
2) Dry
3) Both

**# of nights stayed:**
1) < 1 day
2) 1-2 days
3) > 2 days

**Malaria importation:**
1) Yes
2) No

| **Health Post/Center Characteristics** | | |
|---|---|---|
| Year founded | | |
| Health care workers: (circle one) | | |
| Physicians | Yes | No |
| Nurses | Yes | No |
| Midwives | Yes | No |
| Microscopists | Yes | No |
| Services offered: (circle one) | | |
| Family planning | Yes | No |
| Maternal health | Yes | No |
| Pediatric health | Yes | No |
| Malaria diagnostic | Yes | No |
| Malaria treatment | Yes | No |
| Minor illness/injury | Yes | No |
| Surgery | Yes | No |
| Dental | Yes | No |
| Other | Yes | No |
| **Data Upload to REDCap** | | |
| Uploaded by: | | |
| Date (dd/mm/yy): | | |

| **Community Name:** ____________ | **Reason for travel** | **# of people coming in a year (0-10000)** | **Season** | **Travel time** | **# of nights stayed in a year** | **Malaria importation** | **Notes** |
|---|---|---|---|---|---|---|---|

**Web-scraping Activity**

We conducted a thorough search of routine and singular surveys carried out by the federal governments of Peru, Ecuador, and Brazil available on their respective websites (National Institute for Statistics and Information (INEI), National Institute of Statistics and Censuses (INEC), and Brazilian Institute of Geography and Statistics (IBGE)) as well as data from the most recent censuses. Census data regarding household construction and occupation related data were collected from the Integrated Public Use Microdata Series (IPUMS-I) at the University of Minnesota. Only data available on administrative level three or more granular were included for Ecuador and Peru and administrative level two in Brazil. To normalize the data, proportions and averages were calculated to avoid inflating the values for large administrative areas.

These data were harmonized across the three countries by identifying variables encoding similar data. These variables were re-coded to correspond with the most granular data available common across all three countries. Individual observations were then collapsed to obtain one observation to represent their respective administrative area. National level datasets were created and then consolidated into one file.

The Brazilian data is incomplete at this time due to challenges in comparability with data from Ecuador and Peru and general availability. Much of the data identified for Brazil in the public domain was not granular enough to include in this dataset, being aggregated to administration level one. Further differences exist in questions included in census data and the coding of responses that limited use in the harmonization process.

**Supplementary Table 1.** Summary of data sets compiled from web scraping activity

| Data Category | Description of data | Year of Collection | Method of collapse |
|---|---|---|---|
| Demographic Characteristics | Total population count | Peru: 2017 | Summation |
| | | Ecuador: 2010 | |
| | | Brazil: 2010 | |
| | Count of the individuals by gender (male/female) | Peru: 2017 | Proportion |
| | | Ecuador: 2010 | |
| | | Brazil: 2010 | |
| | Count of the individuals by 5-year age categories | Peru: 2017 | Proportion |
| | | Ecuador: 2010 | |
| | | Brazil: 2010 | |
| Educational Infrastructure | Total number of schools classified as initial (pre-primary school), primary, secondary, and total number of schools in the lowest administrative level for the respective country | Peru: 2013 | Summation |
| | | Ecuador: 2020 | |
| | | Brazil: NF | |
| | Number of students educated at each educational institution | Peru: 2013 | Average |
| | | Ecuador: 2020 | |
| | | Brazil: NF | |
| | If an individual indicated that they attended school outside of their home administrative unit (Parrish, District, Municipality) | Peru: 2017 | Proportion |
| | | Ecuador: 2014 | |
| | | Brazil: NF | |
| Health infrastructure | Level of health posts in the given administrative area in terms of services provided | Peru: 2015 | Summation |
| | | Ecuador: 2018 | |
| | | Brazil: NF | |
| | Total number of health posts irrespective of level in the given administrative level | Peru: 2015 | Summation |
| | | Ecuador: 2018 | |
| | | Brazil: NF | |

| | | | |
|---|---|---|---|
| | Coordinates (latitude and longitude) for health post location | Peru: 2015<br>Ecuador: 2018<br>Brazil: NF | Not collapsed |
| Housing | Main method of access route to the residence (road, river, walking path, etc.) | Peru: 2020<br>Ecuador: 2010<br>Brazil: NF | Proportion |
| | Construction materials used for walls, roofs, and floors of the home | Peru: 2017<br>Ecuador: 2014<br>Brazil: 2010* | Proportion |
| | Identification of sewage system associated with the residence | Peru: 2017<br>Ecuador: 2014<br>Brazil: 2010 | Proportion |
| | Identification of where homes obtain drinking water | Peru: 2017<br>Ecuador: 2014<br>Brazil: 2010 | Proportion |
| | Binary variable for if there is electric lighting at the residence | Peru: 2017<br>Ecuador: 2014<br>Brazil: NF | Proportion |
| Telecommunications | Binary variable for if the household has internet access at the residence | Peru: 2017<br>Ecuador: 2014<br>Brazil: 2010 | Proportion |
| | Binary variable for if the household has at least one cell phone at the residence | Peru: 2017<br>Ecuador: 2014<br>Brazil: 2010 | Proportion |
| | Binary variable for if the household has a fixed phone (landline) at the residence | Peru: 2017<br>Ecuador: 2014<br>Brazil: 2010 | Proportion |
| | Binary variable for if the household has a computer at the residence | Peru: 2017<br>Ecuador: 2014<br>Brazil: 2010 | Proportion |
| Work/Occupation | Number of employees at businesses | Peru: 2015<br>Ecuador: 2014<br>Brazil: NF | Average |
| | Binary variable for in the business has electricity | Peru: 2015<br>Ecuador: 2014<br>Brazil: NF | Proportion |
| | Type of occupation by individual categorized as:<br>• Agriculture, fishing, and forestry<br>• Mining and extraction<br>• Manufacturing<br>• Electricity, gas, water, and waste management<br>• Construction<br>• Wholesale and retail trade<br>• Hotels and restaurants<br>• Transportation, storage, and communications<br>• Financial services and insurance<br>• Public administration and defense<br>• Services, not specified<br>• Business services and real estate<br>• Education<br>• Health and social work<br>• Other services<br>• Private household services<br>• Other industry | Peru: 2007<br>Ecuador: 2014<br>Brazil: 2010 | Proportion |
| | Type of work for each business categorized as: | Peru: 2013 | Proportion |

| | | | |
|---|---|---|---|
| | • Creating goods for market such as weaving, making clothes, furniture making<br>• Prepare and/or selling food<br>• Working on cars or construction work<br>• Fishing, lumber, gold mining industries<br>• Service industry including but not limited to taxi drivers, servers, domestic workers, laundry<br>• Other jobs | Ecuador: 2014<br>Brazil: NF | |
| | Individual's total personal income from all sources in the previous month or year (USD 2022) | Peru: 2021<br>Ecuador: 2014<br>Brazil: 2010 | Average |
| | Average number of hours worked per week over the past 2 weeks to 1 month | Peru: 2021<br>Ecuador: 2022<br>Brazil: 2010 | Average |
| | Binary variable for if the individual works in the same administrative area in which they reside (level three for Peru and Ecuador, level two for Brazil) | Peru: 2017<br>Ecuador: 2014<br>Brazil: NF | Proportion |

* Wall material only
NF – Not found (at time of publication)

### Land Data Assimilation System (LDAS)

NASA's Land Information System (LIS) Land Data Assimilation System (LDAS) software platform incorporates various data sources and models to improve our understanding of land surface processes (1,2). In this application, we draw meteorological forcing for the LDAS simulations from the Global Data Assimilation System (GDAS), supplemented with precipitation estimates from NASA's Global Precipitation Measurement (GPM) Integrated Multi-satellite Retrievals for GPM (IMERG) product. The GDAS data are topographically downscaled and integrated into the system using LIS's Land Surface Data Toolkit (LDT) for a unified spatial resolution over the different years (3). Details of this LDAS implementation are available in (4).

The land surface model employed in this setup is the Noah-Multiparameterization (Noah-MP) model, an enhanced version of the Noah Land Surface Model (LSM). Noah-MP represents hydrologic variables and land surface energy balance through physically based hydrologic processes. It incorporates multiple options for various physical processes and augments the representation of surface energy balance, snow and frozen soil, groundwater, runoff, and leaf dynamics. The Noah-MP model provides water fluxes and energy balance within the land surface.

To initialize the system, a 40-year offline spin-up simulation covering the period 2001-2020 twice is performed to achieve equilibrium hydrological states. These states help establish the best estimation of hydrological outputs when the monitoring period begins. The LDAS runs Noah-MP on a 30-minute time step and generates water and energy fluxes and states at a 5 km spatial resolution, saving the outputs at a daily temporal resolution. The system utilizes satellite-informed input parameters, such as land cover data from the Moderate Resolution Imaging Spectroradiometer-International Geosphere Biosphere Program (MODIS-IGBP), elevation data from the Shuttle Radar Topography Mission (SRTM), and albedo and vegetation fraction maps derived from NOAA's polar orbiting satellites, along with soil texture data from the Food and Agriculture Organization (FAO). This setup has been implemented in retrospective mode from 2001 to 2022, with a restart file from the spin-up simulation used as initialization.

Concurrent with the Noah-MP model, the HyMAP model operates over the same geographical domain to estimate streamflow throughout the river network. Noah-MP and HyMAP are one-way coupled, with Noah-MP runoff feeding into HyMAP, and their combined outputs provide valuable insights into hydrological processes and the overall water cycle (Supplementary Table 2).

**Supplementary Table 2**: List of LDAS output variables generated from the retrospective model run.

| | **LDAS output variable** |
|---|---|
| 1 | Net shortwave radiation |
| 2 | Net longwave radiation |
| 3 | Latent heat flux |
| 4 | Sensible heat flux |
| 5 | Ground heat flux |
| 6 | Snowfall rate |
| 7 | Rainfall rate |
| 8 | Total evapotranspiration |
| 9 | Surface runoff |
| 10 | Subsurface runoff |
| 11 | Snowmelt |
| 12 | Surface albedo |
| 13 | Snow water equivalent |
| 14 | Average layer soil moisture |
| 15 | Average layer soil temperature |
| 16 | Potential evapotranspiration |
| 17 | Interception evaporation |
| 18 | Vegetation transpiration |
| 19 | Bare soil evaporation |
| 20 | Root zone soil moisture |
| 21 | Water table depth |
| 22 | Terrestrial water storage |
| 23 | Groundwater storage |
| 24 | Near surface wind |
| 25 | Near surface air temperature |
| 26 | Near surface specific humidity |
| 27 | Surface pressure |
| 28 | Surface incident shortwave radiation |
| 29 | Surface incident longwave radiation |
| 30 | Leaf area index |
| 31 | Greenness |
| 32 | Streamflow |
| 33 | River depth |
| 34 | Flooded area |
| 35 | River flow velocity |
